# Highly efficient and sensitive membrane-based concentration process allows quantification, surveillance, and sequencing of viruses in large volumes of wastewater

**DOI:** 10.1101/2023.09.25.23296071

**Authors:** G. El soufi, L. Di Jorio, Z. Gerber, N. Cluzel, J. Van Assche, D. Delafoy, R. Olaso, C. Daviaud, T. Loustau, C. Schwartz, D. Trebouet, O. Hernalsteens, V. Marechal, S. Raffestin, D. Rousset, C. Van Lint, JF. Deleuze, M. Boni, OBEPINE consortium, O. Rohr, M. Villain-Gambier, C. Wallet

## Abstract

Wastewater-based epidemiology is experiencing exponential development. Despite undeniable advantages compared to patient-centered approaches (cost, anonymity, survey of large populations without bias, detection of asymptomatic infected peoples…), major technical limitations persist. Among them is the low sensitivity of the current methods used for quantifying and sequencing viral genomes from wastewater. In situations of low viral circulation, during initial stages of viral emergences, or in countries experiencing heavy rains, the extremely low concentrations of viruses in wastewater may fall below the limit of detection of the current methods. The availability and cost of the commercial kits, as well as the requirement of expensive materials, can also present major blocks to the development of wastewater-based epidemiological survey, specifically in low-income countries. Thereby, highly sensitive, low cost and open-access methods are still needed to increase the predictability of the viral emergences, to survey low-circulating viruses and to allow wastewater-based surveillance. Here, we outline and characterize new protocols for concentrating, quantifying, monitoring, and sequencing SARS-CoV-2 from large volumes (500 mL-1L) of raw wastewater. Our nucleic acid extraction technique (the routine C: 5ml method) does not require sophisticated equipment such as automatons and is not reliant on commercial kits, making it readily available to a broader range of laboratories for routine epidemiological survey. Furthermore, we demonstrate the efficiency, the repeatability, and the high sensitivity of a new membrane-based concentration method (MBC: 500 mL method) for enveloped (SARS-CoV-2) and naked (FRNAPH GGII) viruses. We show that the MBC method allows the quantification and the monitoring of viruses in wastewater with a significantly improved sensitivity. In contexts of low viral circulation, we report quantifications of SARS-CoV-2 in wastewater at concentrations below 100 genome copies per liter and as low as 40 genome copies per liter. In highly diluted samples collected in wastewater treatment plants of French Guiana, we confirmed the accuracy of the MBC method compared to the estimations done with the C method. Finally, we demonstrate that both the C and MBC methods are compatible with SARS-CoV-2 sequencing. We show that the quality of the sequence is correlated with the concentration of the extracted viral genome. Of note, the quality of the sequences obtained with some MBC processed wastewater was improved by dilutions or enzyme substitutions suggesting the presence of specific enzyme inhibitors in some wastewater. To the best of our knowledge, our MBC method is the first efficient, sensitive, repeatable, and up-scalable method characterized for SARS-CoV-2 quantification and sequencing from large volumes of wastewater.

**Graphical Abstract:** 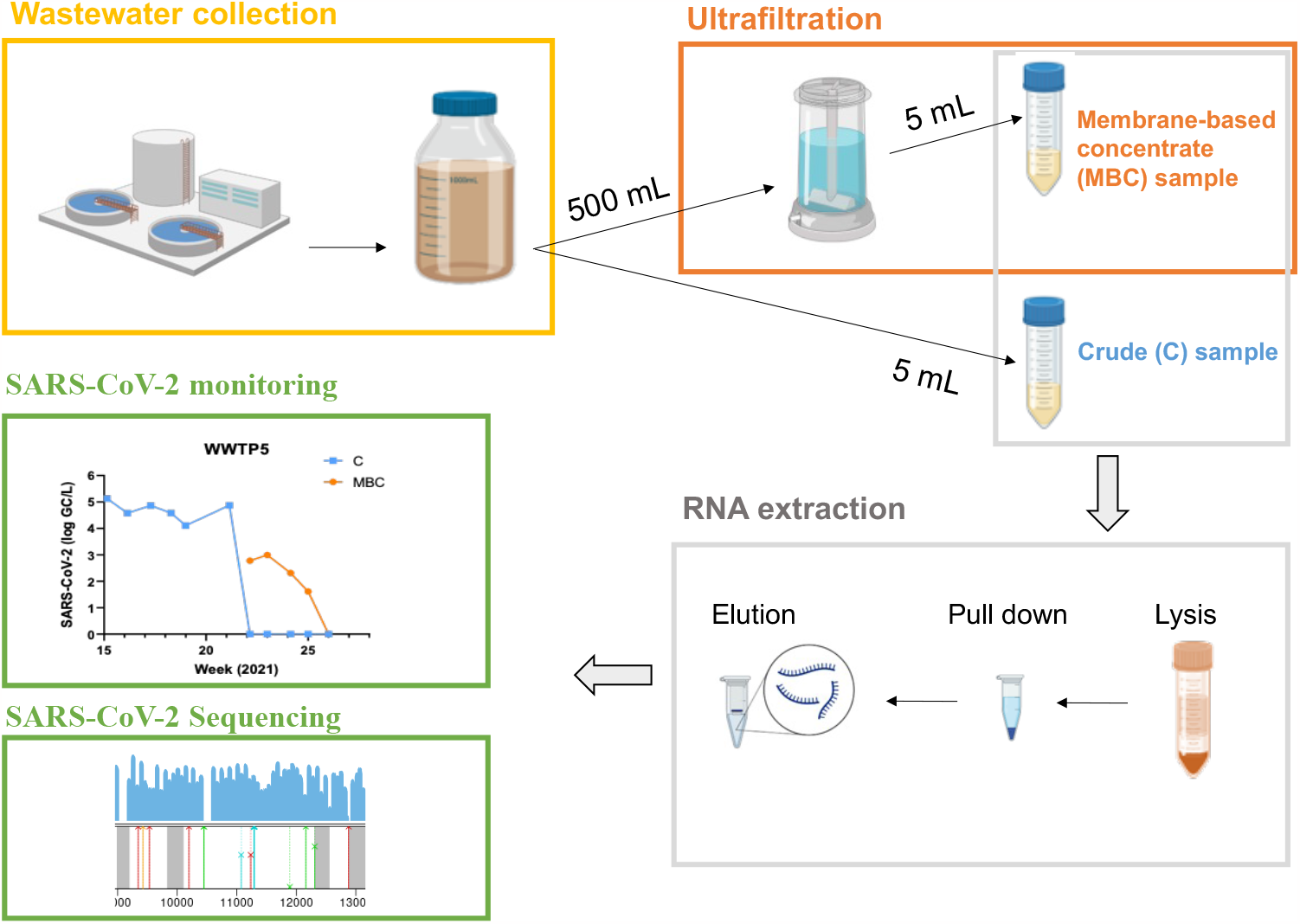

## 1. Introduction

In December 2019, an outbreak of a novel coronavirus (SARS-CoV-2) was reported in China, and quickly spread worldwide. Since then, the ensuing disease (COVID-19) has become a pandemic that has led to several million deaths globally as of February 2023 (WHO). It has become evident that a quick and thorough assessment of the dynamics of SARS-CoV-2/COVID-19 infection within a population is crucial for implementing effective public health actions in response to the pandemic.

The time tested Wastewater-Based Epidemiology (WBE) which has been used to track the lifespan of previous pathogenic viruses such as poliovirus, enterovirus, adenoviruses and noroviruses in communities around the world (Lago et al., 2003, Katayama et al., 2008, and McCall et al., 2020), proved to be equally effective to track SARS-CoV-2. Fecal shedding of SARS-CoV-2 with detectable viral RNA found in domestic waters has been reported and demonstrated by Kitajima et al., 2020, and Xiao et al., 2020. Furthermore, recent studies have identified a correlation between the number of viral genome copies in wastewater and the prevalence of infection within the population (Ahmed et al., 2020a; Medema et al., 2020a; Wurtzer et al., 2020). As such, WBE has been successfully adapted as a public health tool for tracking the spread of SARS-CoV-2 within communities (Maréchal et al., 2023). Worldwide Public Health departments have been diligently researching rapid and cost-effective tools for monitoring SARS-CoV-2 in wastewater samples. Regrettably, most protocols used for tracking SARS-CoV-2 in wastewater rely on viral extractions from small sample volumes. These protocols lack the ability to scale-up the testing capacity (Zheng et al., 2023). Consequently, when the virus is present at low levels (during epidemic slowdowns, or due to wastewater dilution from rainfall, for exemple), these protocols demonstrate reduced accuracy and therefore cannot achieve sufficient sensitivity (Zheng et al., 2023). Thus, there is a need for a more sensitive protocol for detecting and quantifying the virus. This protocol should have the capacity to detect SARS-CoV-2 RNA when the number of cases is low and should be applicable to monitor the emergence of specific variants.

Prior to this work, various virus concentration methods were developed and used by scientists worldwide. These methods included ultrafiltration, polyethylene glycol (PEG) precipitation, direct sludge extraction, and skimmed milk flocculation (Ahmed et al., 2020, Peccia et al., 2020, Medema et al., 2020). However, there is no documented successful optimization for enveloped and naked viruses concentration from large wastewater volumes like 500mL and more.

An extensive study by (Ahmed et al., 2020b) compared seven viral concentration methods for the RT-qPCR-based recovery of murine hepatitis virus (MHV), a surrogate for SARS-CoV-2 in untreated wastewater. Most of these methods required expensive instruments. For example, the adsorption-extraction method using an electronegative membrane required ideally a bead-beating system (Ahmed et al., 2020b) . The ultracentrifugation method required expensive equipment (ultracentrifuge) which may not be available in routine analysis laboratories (Ahmed, Bertsch, et al., 2020). Moreover, most reported concentration methods were tailored for small volumes of wastewater samples. For example, the centrifugal filter devices like Amicon Ultra15 (30kDa) can process only 15 mL of sample at a time. With the ultracentrifugation method, samples volumes could be processed up to 50 mL at a time. In a recent study, Zheng et al., 2023 has developed a PEG-precipitation method for SARS-CoV-2 wastewater surveillance but the volume processed was again limited to 40 mL. Mc Minn et al. described a concentration method applicable for 2L of wastewater (McMinn et al., 2021). However, the concentration process requested quite expensive instruments, excluded the solid compartment of the wastewater, demonstrated a limited recovery rate around 20%, and was not evaluated for genome sequencing. Hence, there is an urgent need for a deeply characterized, simple, sensitive, and affordable new virus concentration method for large-scale surveillance.

In this study, we implemented new protocols for the concentration, quantification, monitoring, and sequencing of SARS-CoV-2 from large (500 mL-1L) raw wastewater volumes. The samples were collected from four wastewater treatment plants (WWTPs) in the Alsace region, France and in two WWTPs in French Guiana. Our open access, efficient, sensitive, and low-cost Membrane-Based Concentration (MBC) process allows efficient concentrations, quantifications, and sequencing of SARS-CoV-2. Our nucleic acid extraction method for crude wastewater samples (routine C method) and MBC processed samples, unlike others, does not rely on commercial kits, making it readily accessible to a wider range of laboratories. Moreover, it does not require sophisticated equipment such as automatons; all steps can be done manually. Furthermore, our new protocols allow as well efficient concentration and quantification of FRNAPH GGII, a non-enveloped virus. In this article, we demonstrate the efficiency, the repeatability, and the sensitivity of our new ultrafiltration-based concentration method for both enveloped (SARS-CoV-2) and naked (FRNAPH GGII) viruses. We show that our large volume-based method allows for the quantification and the survey of viruses in wastewater with significantly improved sensitivity. We report SARS-CoV-2 quantifications below 100 genome copies per liter and even as low as 40 genome copies per liter. Finally, we show that our methods are compatible with sequencing. To the best of our knowledge, our MBC method is the first efficient, sensitive, repeatable, and up-scalable method described for SARS-CoV-2 quantification and sequencing from large volumes of wastewater.

## 2. Material and methods

### 2.1. Sample collection

Samples of 24-h-composite raw wastewater (influent) were collected weekly from Alsace region (France) and French Guiana wastewater treatment plants. Samples were shipped to the laboratory under cool conditions, stored at 4 °C, and used within one week of collection. Information on sampling points in Alsace is shown in Supplementary Table 1.

The conductivity was measured using Protavo® 907 MULTI (SE 615/1-MS, Knick), the pH was measured using a pH meter Accumet AE150 (Fisher Scientific) and the optical density (OD) at 600 nm was measured using an Eppendorf Biophotometer.

### 2.2. Viral concentration method (MBC method)

An aliquot of 500 mL of each wastewater sample was decanted for 30 min in order to sediment suspended solids that may interfere with the ultrafiltration. The aliquot was concentrated using an Amicon® Stirred Cell 400ML (UFSC40001, Merck-Millipore), with a cut-off of 10 kDa membrane (Ultracel® regenerated cellulose membrane, PLGC07610, Merck-Millipore), under a vacuum pressure set at 3 bar. The Ultracel® membrane was pre-rinsed before use, following manufacturer instructions, and then the aliquot of 500 mL was filtered, starting with the supernatant and pouring the sediments at the end, to obtain the final concentrate of 5 mL. The membrane was rinsed by adding 4.5 mL of lysis buffer pH 4 (Guanidine Thiocyanate 10M / Tris-HCl 0.1M pH 6.4, 2% SDS, EDTA 0.038 M) into the cell. The lysis buffer was collected and added to the membrane-based concentrate sample (MBC).

### 2.3. Nucleic acid extraction (routine C method)

Viral nucleic acids of crude samples (C) and membrane-based concentrate samples (MBC) were extracted using the protocol developed hereafter. The volume of the samples used for the extraction was 5 mL and the elution volumes were 50 μL for C samples and 100 μL (1^st^ elution) and 50 μL (2^nd^ elution) for MBC samples. Negative control of the viral nucleic acid extraction was added per batch of samples to account for any contamination during extraction. 4.5 mL of lysis buffer (Guanidine Thiocyanate 10M / Tris-HCl 0.1M pH 6.4, 2% SDS, EDTA 0.038 M) and 450 μL of 2% DTT were added to each sample. The samples were incubated for 10 min at room temperature and then for 10 min at 60°C.

A first step of purification with acid phenol pH 4.3 / chloroform 1:1 (v/v) was carried out for MBC samples and C samples dedicated to comparisons with MBC samples but not for C samples dedicated to routine surveillance. A volume of 9.5 mL of phenol / chloroform mixture was added to each sample. Grease tubes (silicone grease / silica 90:10 (w/w)) were used to facilitate phase separation. The samples were centrifuged for 5 min at 3500 x g and the supernatant was collected.

For the crude wastewater samples processed with the routine C method (no phenol/chloroform step needed) and the MBC processed samples, 15 μL of polyadenylic acid (10 g/L), 5 mL of isopropanol, and 8 μL of silica particles (1.04 g/cm3) (MagPrep® Silica Particles, Sigma-Aldrich) were next added. Samples were incubated for 10 min with shaking at room temperature. Then, the beads were collected and then washed twice with a washing buffer (lysis buffer / ethanol 1:1 (v/v) and DTT 2%) and twice with another buffer (ethanol / Nuclease-Free Water 8:2 (v/v)). Next, the beads were incubated for 10 min at room temperature. Elution was carried out for samples C by adding 50 μL of elution buffer (Tris-HCl-EDTA pH9 1M), then the sample was incubated for 10 min at 95°C. For membrane-based concentrate samples (MBC), two elution steps were performed by adding first 100 μL of elution buffer then 50 μL. A final purification step was performed for MBC samples using the OneStep-96 PCR Inhibitor Removal kit (#ZD6035, OZYME) following manufacturer instructions. This final step was not essential for the viral extraction (results not shown in this paper) but performed to improve the quantification and sequencing processes.

### 2.4. Reverse Transcription -Quantitative Polymerase Chain Reaction (RT-qPCR)

One-step RT-qPCR was performed with Master Mix Fast Virus 1-Step TaqMan™ kit (#4444434, Applied Biosystems – ThermoFischer Scientific) on a BioRad CFX96™ thermal cycler, software version 3.1 (Bio-Rad Laboratories) manually setting the threshold and baseline. Two RT-qPCR assays were chosen to quantify SARS-CoV-2 : (I) E-Sarbeco assay (Corman et al., 2020) targeting the envelope protein E and (II) RdRp-IP4 assay targeting part of the ORF1ab. Primers and probes were synthesized and provided by Eurofins Genomics (sequences available in supplementary table 2). nCoV_E-Sarbeco and nCoV_IP4 set were used with 0.4 μM of each primer, 0.2 μM of probe in 10 μL final reaction volume with 5 μL of RNA sample, and 2.5 μL of Master Mix Fast Virus 1-Step TaqMan™ for each assay.

The presence of PCR inhibitor in RNA extracts was assessed using a dengue RT-qPCR assay after spiking a known copy number of Dengue RNA (exogenous RNA, which is not present in the wastewater samples). In order to determine PCR inhibition, Dengue RNA was also added to Nuclease-Free Water sample and the resulting Ct value was used to set-up a reference point. Subsequently, Dengue RT-qPCR assay was performed in 10 μL reaction mixtures using Bio-Rad CFX96 thermal cycler (Bio-Rad Laboratories), manually setting the threshold and baseline. Dengue set (Euliano et al., 2019) (sequences available in supplementary table 2) was used with 0.4 μM of each primer, 0.2 μM of probe in 10 μL final reaction volume with 5 μL of RNA sample and 2.5 μL of Master Mix Fast Virus 1-Step TaqMan™ and 6.8 x 10^3^ genome copies / reaction of Dengue RNA. All samples were analyzed alongside two no template controls. All wastewater samples were within the 2-Ct values of the reference Ct.

Furthermore, F-specific RNA bacteriophages were quantified in all wastewater samples to determine the efficiency of viral genomes concentration. Briefly, for the FRNAPH GGII genome, the VTB4-Fph GII set published by Wolf et al., 2010 was used with 0.4 μM of each primer, 0.2 μM of probe in 10 μL final reaction volume with 2.5 μL of RNA and 2.5 μL of Master Mix Fast Virus 1-Step TaqMan™. On the BioRad thermal cycler, the RT was conducted at 50°C for 5 min followed by an initial denaturation cycle at 95°C for 20s and 49 cycles including denaturation step at 95°C for 3s, and hybridization step at 58°C for 30s.

### 2.5. LOD/LOQ determination

Synthetic quantified SARS-CoV-2 RNA provided by Biorad (ref COV019) was used. A standard curve was prepared and ran in 2 replicates in a fourfold dilution series with concentrations at 1 to 1000 genome copies per reaction. A Ct ≥ 40 was considered as negative. The lowest concentration that produced at least 95% positive replicates was assumed to be the limit of detection (LoD) of the RT-qPCR. The limit of quantification (LoQ) was estimated by runs test following linear regression. Values were excluded of regression until linearity was reached.

### 2.6. Viral recovery efficiency

Process recovery efficiency represents an important metric in viral signal quantification, as it allows comparison of results from study to study even if different concentration and quantification methods are used. In this study, the efficiency of virus recovery from the whole process (ultrafiltration / RNA extraction / RT-qPCR) was calculated using the following equation;

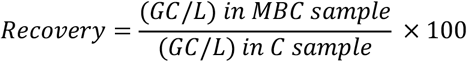

### 2.7. SARS-CoV-2 RNA sequencing

NGS sequencing libraries were prepared using COVIDSeq Test kit (Illumina, Inc San Diego, CA, USA) with SARS-CoV-2 specific primers Artic v4.1 as described in (Gerber et al., 2022). The SARS-CoV-2 enriched libraries were sequenced using NovaSeq 6000 SP Reagent Kit v1.5 (300 cycles) according to the manufacturer’s instructions (Illumina, Inc San Diego, CA, USA). Sequencing data was analyzed using our in-house bioinformatic pipeline as described in (Gerber et al., 2022) using Pangolin database v4.2 (O’Toole et al., 2021).

### 2.8. Statistical analysis

All statistical analyses were performed with GraphPad Prism version 9 as indicated in the figures. Differences between groups were considered significant when the *P* value < 0.05.

## 3. Results

### 3.1. Membrane-based concentration process is efficient to enrich enveloped and naked viruses from large volumes of wastewater

Concentrating viruses from wastewater samples is a crucial step, especially for detecting low-level virus RNA during the onset, the resurgence, and the deceleration of epidemics or in diluted wastewater due to rainfall. Twenty-three wastewater samples were collected from four different WWTPs in Alsace, France from February to October 2022 and were subsequently analyzed. Half-liter of each wastewater was concentrated using ultrafiltration (Amicon® 10 kDa) to yield a final concentrate of 5 mL (MBC for membrane-based concentration). Viral RNA was then extracted using our in-house protocol and quantified by analyzing specific genes E-Sarbeco and RdRp-IP4 for SARS-CoV-2 and VTB4-Fph GII for FRNAPH GGII, using RT-qPCR. RNA amounts in membrane-based concentrated (MBC) samples were compared to their corresponding crude wastewater samples (C for crude) (Figure 1).

**Fig. 1:**
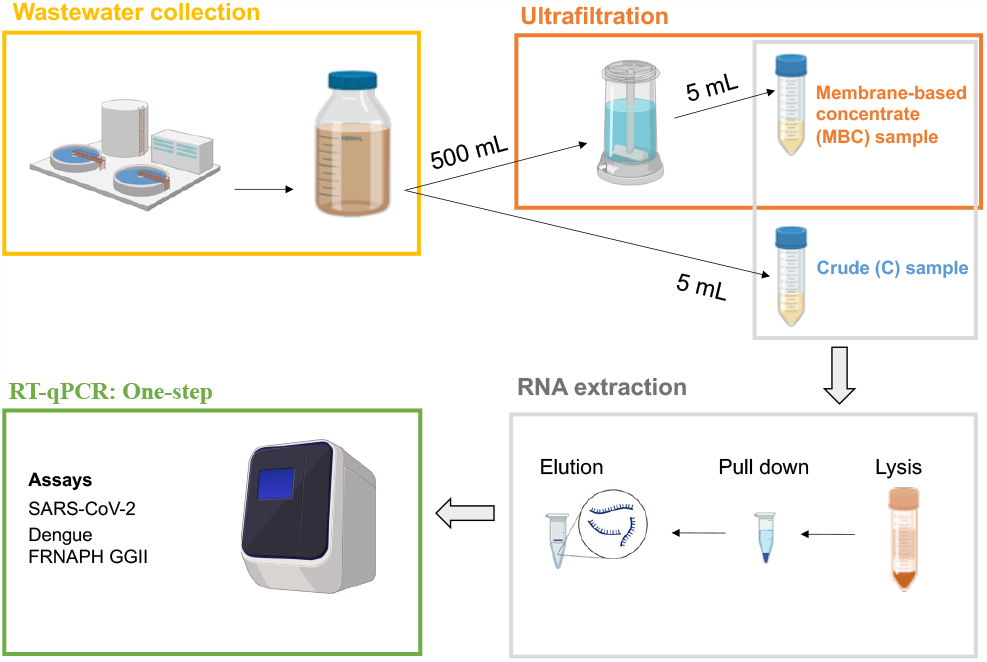
Overview of the process used for quantifying SARS-CoV-2 in crude (C) and membrane-based concentrated (MBC) large volume of wastewater. 24h representative wastewater samples were collected from four different wastewater treatment plants (WWTPs) across Alsace, France. The MBC samples were concentrated from 500ml to 5ml through ultrafiltration using Amicon® Stirred Cell 400ML and Ultracel® regenerated cellulose membrane (10 kDa). Crude and MBC samples were subjected to RNA extraction and the RT-qPCR quantifications.

As shown in figure 2A, the extracted amount of SARS-CoV-2 RNA, measured in genome copies (GC), ranged from 5.9 x 10^1^ to 5.5 x 10^3^ and from 3.8 x 10^3^ to 2.5 x 10^5^ for C and MBC samples, respectively. The process allowed an average increase of 51 ± 24-fold in SARS-CoV-2 quantities in MBC samples compared to C samples. Notably, some samples were enriched up to 97-fold, approaching the technique’s 100% efficiency for the input of 500 ml. The theoretical limit of detection corresponding to 1 GC per reaction is 2.0 x 10^3^ GC/L for 5ml of crude wastewater (C method). Based on our standard curves, the measured limit of detection (95% of detection rate) for C samples was below 3.0 x 10^4^ genome copies per liter (GC/L), with detections possible even below 7.8 x 10^3^ GC/L (Supplementary Fig 1 and 2). However, in the context of the weekly epidemiological monitoring of 11 wastewater treatment plans in Alsace-France, we often measured concentrations around 2000 GC/L suggesting the technique’s limit of detection closely align with the theoretical one. Moreover, comparisons with a commercial 5mL-based method confirmed that our open-access routine C method is suitable for SARS-CoV-2 monitoring (Supplementary fig 3A protocol 1 vs protocol 2 and Supplementary fig 3B). The high efficiency of the MBC process allowed for quantifications below 125 and up to 40 GC/L. As shown in supplementary figure 3A protocol 3, MBC process proves valuable for quantifying SARS-CoV2 when the concentrations in wastewater fall below the LOD of the routine C method. Notably, this high sensitivity of the MBC process was reachable for input volumes of 500 ml and 1L. However, MBC is suitable for larger volumes, potentially enabling quantification of even lower quantities of the virus.

**Fig. 2:**
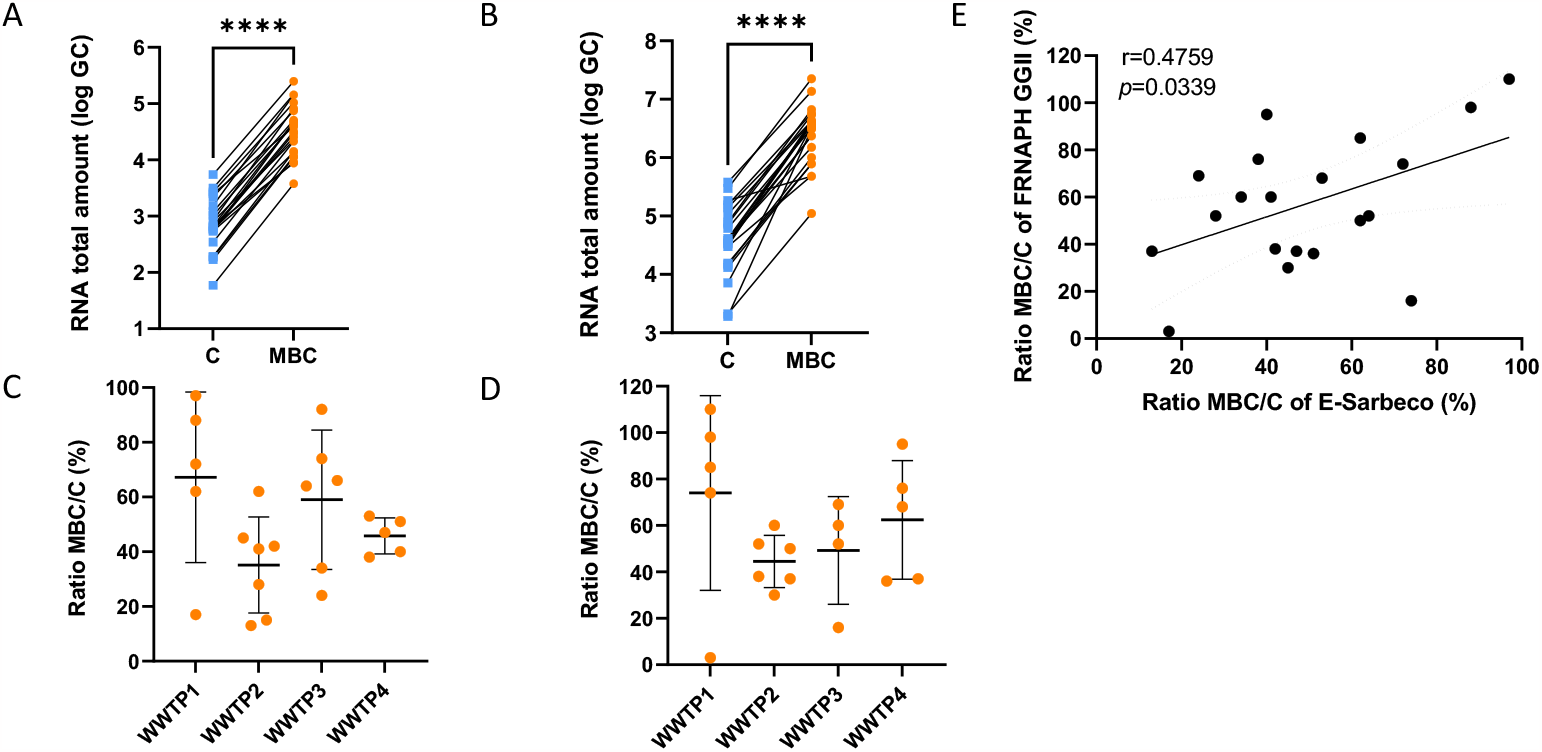
Membrane Based Concentration (MBC) is efficient for enveloped (SARS-CoV-2) and naked (FRNAPH GGII) viruses. Total amount of SARS-CoV-2 (target gene E-Sarbeco) (A) and FRNAPH GGII (target gene VTB4-Fph GII) (B) genomes were quantified in 5ml crude (blue) and 500ml Membrane-based concentrated (orange) wastewater samples collected at different time points in four different treatment plants. Wilcoxon test were performed for statistical analysis. **** p<0.0001. Recovery (mean ± SD) of SARS-CoV-2 RNA (C) and FRNAPH GGII (D) from four wastewater treatment plants (WWTPs) using MBC. Means and standard deviations are shown on theses figures. Correlation between recoveries of SARS-CoV-2 and FRNAPH GGII in wastewater samples concentrated by ultrafiltration (E). Pearson’s correlation coefficient (r) and the corresponding p-value are shown on this figure.

To determine the efficiency of the MBC method for naked viruses, FRNAPH GGII RNA was concentrated and extracted using the same method. The extracted amount of FRNAPH GGII RNA ranged from 1.9 x 10^3^ to 3.8 x 10^5^ and 1.1 x 10^5^ to 2.3 x 10^7^ GC/L for C and MBC processed samples, respectively. The average increase of FRNAPH GGII in MBC samples compared to C samples was 57 ± 28-fold (Fig.2B). Since the MBC efficiencies measured for FRNAPH GGII are comparable to those observed for SARS-CoV-2, the lowest detectable quantities are in the same range and below 100 GC/L.

The viral recovery rate was calculated as the ratio of the targeted virus (SARS-CoV-2 and FRNAPH GGII) concentration in MBC samples to its concentration in C samples. The mean recovery efficiency was 51 ± 24% and 57 ± 28%, for SARS-CoV-2 and FRNAPH GGII, respectively, using ultrafiltration method across samples from 4 WWTPs collected at different times between February and October 2022. However, we observed variations in the efficiency of viral RNA recovery between the different collection weeks for each WWTP and between the different WWTPs (Fig 2C and 2D). The standard deviation S.D. of recovery efficiency within WWTPs ranged from 6.6 to 31.2% and from 11 to 42% for SARS-CoV-2 and FRNAPH GGII, respectively (Fig 2C and 2D). These results suggest that the efficiencies of viral enrichment depend on the WWTP being sampled and on the week of collection. The heterogeneity of the recovery rates is therefore linked to the heterogeneity of the wastewater composition. Finding the key chemical and biochemical components that influence MBC efficiencies will need more specific investigations. However, despite the heterogeneity of the wastewater, the overall efficiencies of our MBC process were comparable to those described for smaller volumes of wastewater (Bertrand et al., 2021) and allowed us to quantify very low concentrations of viruses. Of note, MBC is easy to up-scale to further gain sensitivity if needed.

Next, we compared the MBC process for SARS-CoV-2 and FRNAPH GII. As shown in figure 2E, a good correlation could be found between the two viral concentrations after the MBC process. These results suggest that FRNAPH GII can be used as a good internal control for normalization of the SARS-CoV2 concentrations when parameters such as the flow at the entry of the WWTP are missing. In addition, it allows to take into consideration the efficiency of the MBC process when the SARS-CoV-2 is undetectable with the routine C method.

### 3.2. MBC does not significantly increase PCR inhibition

The presence of PCR inhibitors in wastewater is detrimental to the efficient and sensitive quantification of viruses. To ensure that PCR inhibitors are not concentrated along with the viruses during the MBC process, PCR inhibitions in both C and MBC wastewater samples were assessed. This assessment was carried out using a Dengue RT-qPCR quantification, after spiking RNA samples with a known copy number of Dengue RNA. Additionally, Dengue RNA was added to a nuclease-free water sample to establish a reference point for Ct values. An analysis of 23 C and MBC samples showed slight increases, but no significant PCR inhibitions were observed in the MBC samples (Fig 3A). It is noteworthy that the quantified PCR inhibitions in C and MBC samples were far below the commonly acceptable thresholds for wastewater (ΔCt=2 or 78%). These findings suggest that our experimental conditions were suitable for viral genome quantifications in both crude (C) and membrane-based concentrated (MBC) wastewater samples.

**Fig. 3:**
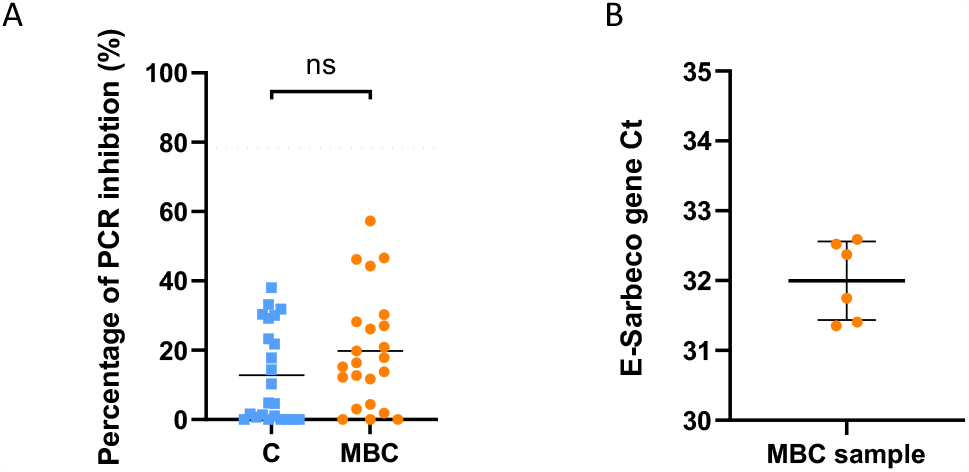
Membrane-based concentration does not significantly increase PCR inhibitors and is highly repeatable. (A) PCR inhibitions have been compared for the crude wastewater (blue) and the membrane-based concentrate samples (orange). A Wilcoxon test has been performed to compare the two groups of samples. ns: not significant. (B) Six different Membrane-based concentrations were performed on the same wastewater and subjected to SARS-CoV-2 E-Sarbeco gene quantifications. Mean and standard deviation are shown on this figure.

### 3.3. MBC is highly repeatable

To determine the intra-assay precision of the entire SARS-CoV-2 quantification process, from the MBC to the final qPCR (ultrafiltration, RNA extraction and RT-qPCR) a wastewater sample collected in week 11 of 2022 from WWTP4 was separately analyzed six times. The Ct values of the SARS-CoV-2 E-Sarbeco gene from these six analyses were plotted against their corresponding mean (Fig. 3B). The relative standard deviation (RSD) to the mean was 1.8%. This RSD of 1.8% is far below the acceptable limit of 15%, as described by Biopharmaceutics Coordinating Committee of the US Food and Drug Administration (Bioanalytical Method Validation. Guidance for Industry, 2018). It is important to note that the RSD to the mean of the extraction part (C method) of the process alone was 1.5% (supplementary Fig. 2). These results demonstrate the high repeatability of our SARS-CoV-2 concentration, extraction, and quantification pipeline. Furthermore, they confirm that the variations in the recovery rate observed in figure 2C and D, were not linked to technical issues but to the heterogeneity of the wastewater.

### 3.4. MBC is suitable for epidemiological survey in wastewater

We next compared the dynamics of viral spread obtained with and without MBC in wastewater from three different WWTPs. SARS-CoV-2 (Fig 4A, B, C) and FRNAPH GGII (Fig 4D, E, F) genome copies per liter of crude wastewater quantified with both C and MBC methods followed the same trend over time. In addition, since the results obtained with our C method processing 5mL were comparable to the one obtained with the commercial NucliSens® EasyMAG™ method for SARS-CoV-2 monitoring in wastewater, the MBC method processing 500mL allowed for wastewater-based epidemiological survey when the quantities of virus fell below the limit of detection of the C method (supplementary Figure 3A protocol 3).

**Fig. 4:**
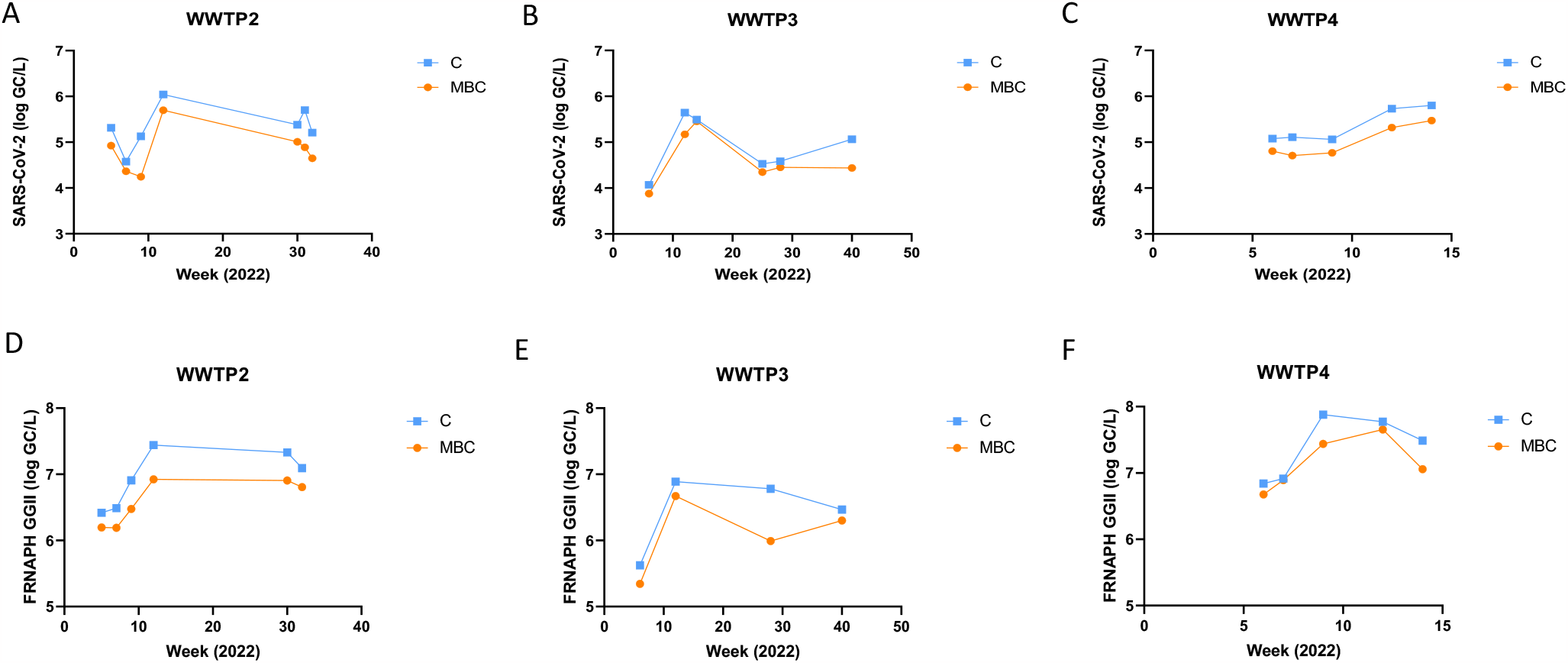
Viral dynamic profiles are similar in crude and membrane-based concentrations. Longitudinal comparison of SARS-CoV-2 (A,B,C) and FRNAPH GGII concentrations (D,E,F) (log genome copies per liter of wastewater) in C and MBC samples . Crude wastewater samples (routine C method) are represented in blue, and the membrane-based concentrated (MBC) samples represented in orange.

To further document this point, we subsequently used wastewater samples collected between April and July 2021 in WWTP2 to perform MBC assays when SARS-CoV-2 concentrations dropped below the detection limit of the C method. As shown in figure 5A, when the virus was no longer detectable in crude samples, MBC allowed the quantification of SARS-CoV-2 for an additional 4 weeks. Thanks to MBC, we were able to quantify up to 40 GC/L of SARS-CoV-2 with a sample input of 1 L. This high sensitivity of the MBC process aligns perfectly with the calculated detection range achievable with 500 mL or 1L, as mentioned in section 3.1. To further test the MBC process in highly diluted wastewater, we quantified SARS-CoV2 in wastewaters samples taken during the transition between the dry and the wet periods in Kourou and Saint Laurent du Maroni (French Guiana). As shown in figure 5B, the concentrations obtained with the C method were close to the LOD and therefore only estimated. The MBC process enable a more precise quantification of these low SARS-CoV2 concentrations confirming that concentrations in proximity to the LOD, as measured with the C method, were at times either overestimated (SLM) or underestimated (Kourou). Of note, SARS-CoV-2 MBC concentrations are presented relative to the ratio GII MBC / GII C to take into consideration the efficiency of the MBC process.

**Fig. 5:**
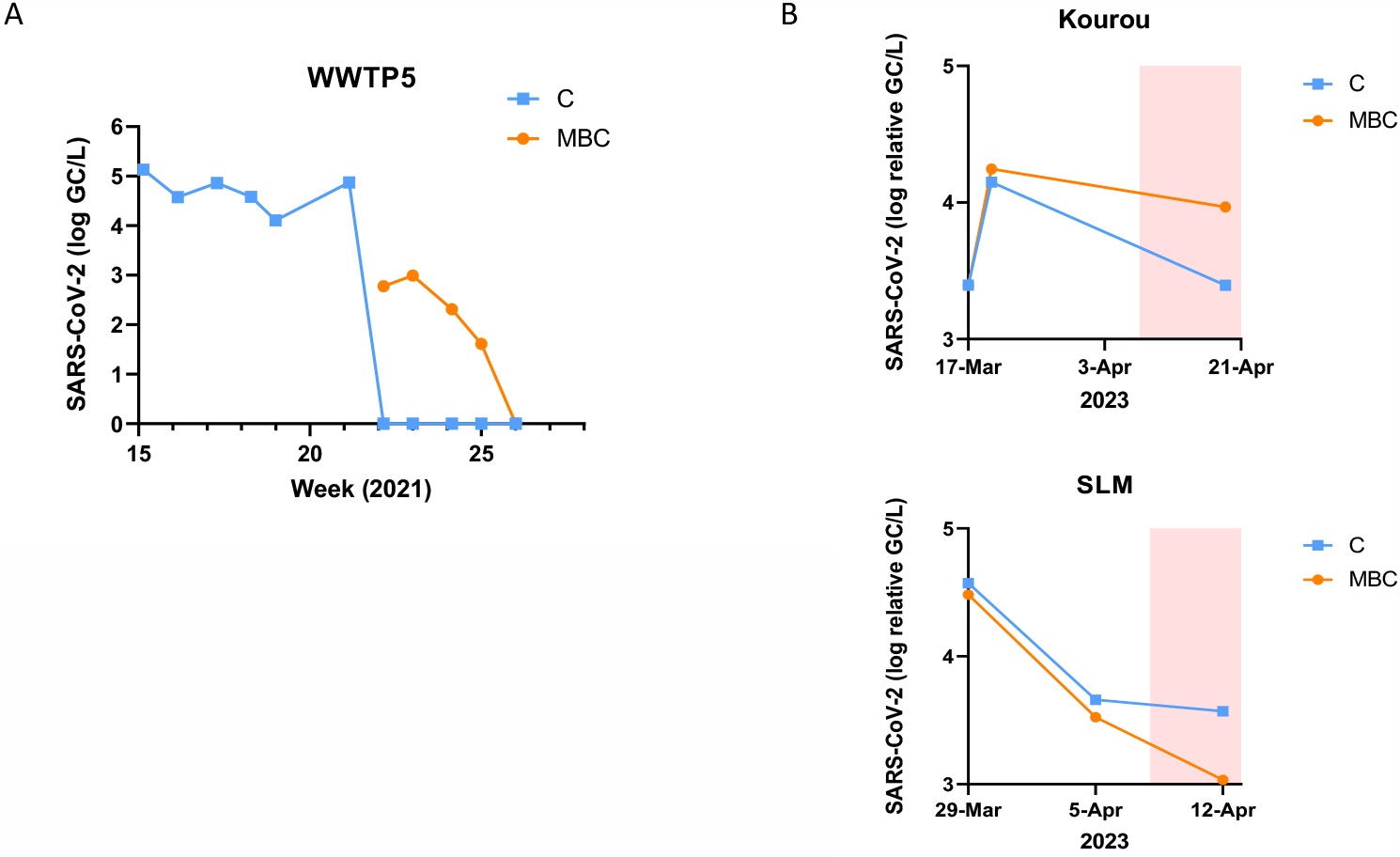
Membrane Based-concentration allows quantifications of low viral concentrations. **(A)** Longitudinal quantifications of SARS-CoV-2 concentration in 5ml crude (blue) and 500ml MBC wastewater from WWTP5. Results are presented in log genome copies per liter. (B) Quantification of Sars-CoV2 in two different wastewater from Guyana before and after the start of monsoon (red aera). GII quantities in the C samples where set to 1 and MBC quantities of SARS-CoV-2 are presented in log GC/L relative to the ratio GII-MBC / GII-C.

### 3.5. MBC allows Next Generation Sequencing of SARS-CoV-2

To investigate the impact of wastewater concentration on Next-Generation Sequencing (NGS), samples of increasing SARS-CoV-2 concentrations were sequenced with the Illumina short reads method COVIDSeq. As expected, the depth of SARS-CoV-2 RNA sequencing in crude wastewater was correlated with the concentration of the samples and close to 100% for concentrations over 20 GC/μl in the final 50μl samples obtained after extraction of 5ml with the routine C method (Fig 6A). This optimal sequencing concentration of SARS-CoV-2 RNA in the extracted sample corresponds to 2.0 x 10^5^ GC/L in the collected wastewater that has been observed only in high epidemic periods and only in 15% of the weekly based-quantified wastewater samples from 11 WWTPs in 2022. These results suggest that MBC could improve the sequencing when the viral concentrations in wastewater are low. We indeed confirm that MBC samples were compatible with NGS but we also observed that some MBC processed wastewater samples were difficult to sequence. These results further demonstrate that the heterogeneity of the origins and the compositions of the samples tested influence the quality of the sequence obtained. However, the dilution of those samples improved the depth of sequencing, suggesting that some NGS enzymes inhibitors present in MBC samples may impact the sequencing efficiency (Fig 6B). Interestingly, the sequencing depth of the diluted MBC samples were better than the one observed with the crude extract of the same wastewater. These results suggest that the PCR inhibitors that did not significantly impact the RT-qPCR step of our process of quantification disfavored NGS sequencing with the COVIDSeq kit. To further investigate this point, we performed NGS sequencing with the full COVIDSeq kit or after substitution of the COVIDSeq reverse transcriptase by the superscript IV reverse transcriptase. Bioinformatic analyses using Pangolin pipeline demonstrated that sequences obtained with MBC samples allow the identification of mutations in the SARS-CoV-2 circulating variant (Figure 7).

**Fig. 6:**
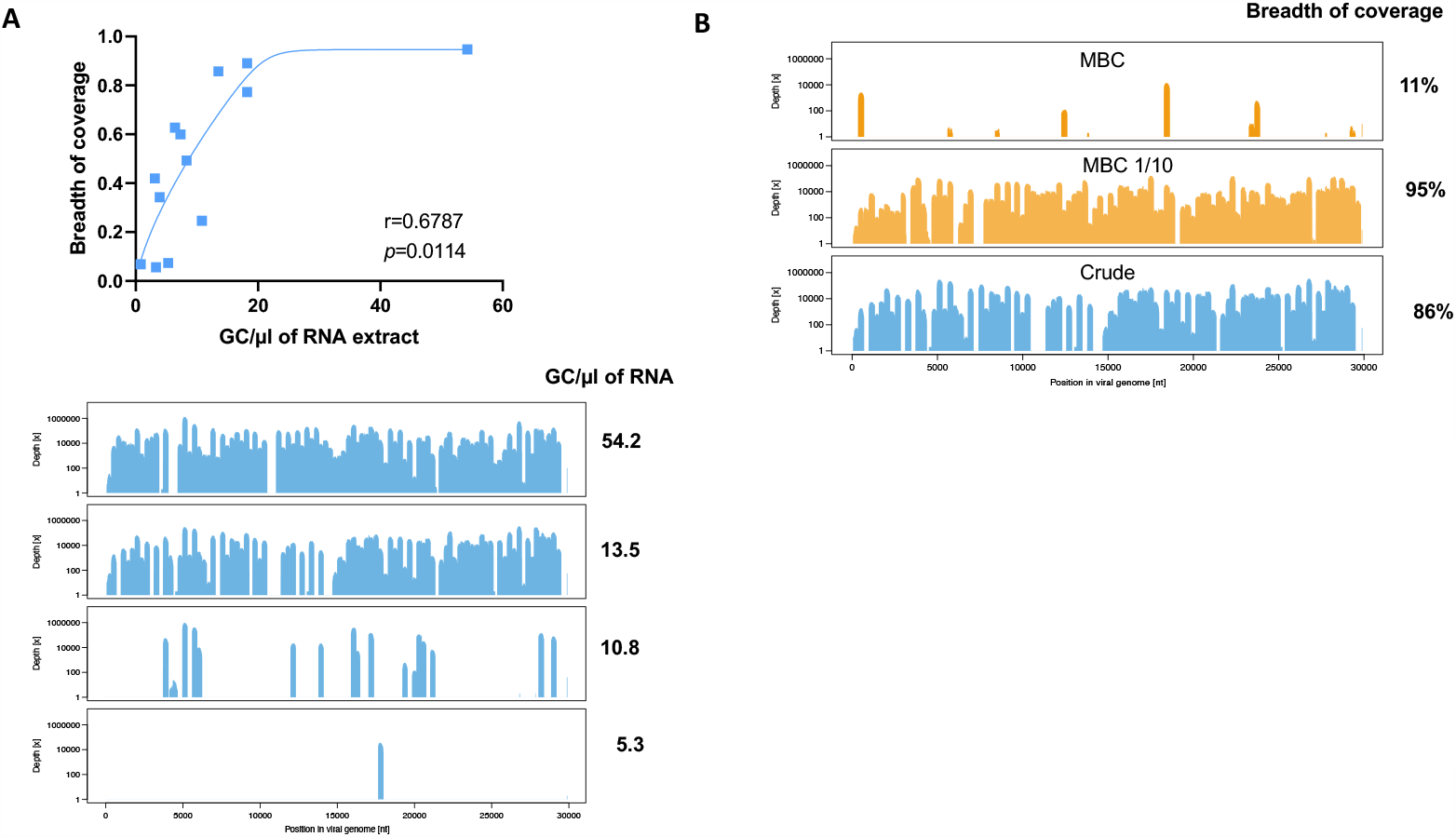
Efficiency of wastewater extracted SARS-CoV2 genome sequencing depends on RNA concentrations (A) and enzymatic inhibitions (B). (A) RNA extracted from different crude wastewater were quantified and subjected to COVID-seq Next-Generation Sequencing. Spearman’s rank correlation results are presented (B) Crude (blue) and MBC (orange) RNA extracts where subjected to COVID-seq Next-Generation Sequencing.

**Fig. 7:**
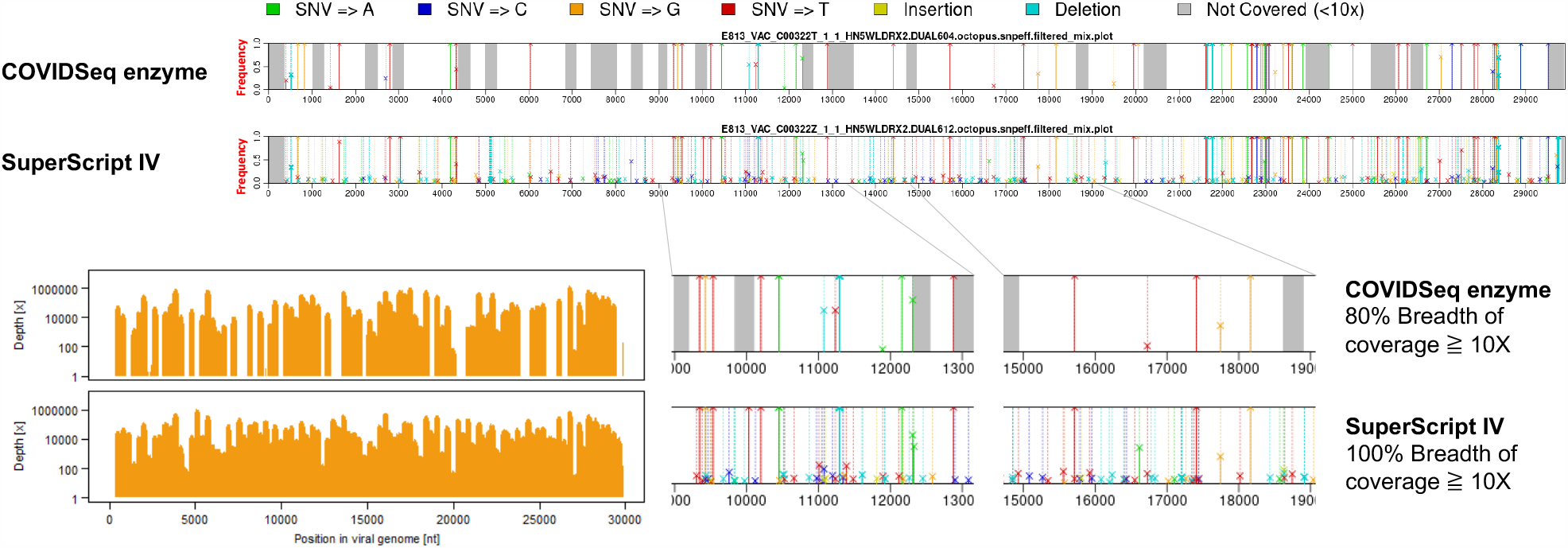
Membrane based concentrated samples are compatible for Next Generation sequencing. MBC RNA extracts were subjected to COVIDseq Next-Generation Sequencing with the complete COVIDSeq kit or with substitutions of the reverse transcription enzyme by SuperScript IV.

Since SuperScript IV improved the depth of sequencing, variant calling highlighted more mutations present at low frequencies. Thereby, the gain of sequence coverage observed with the SuperScript IV was mitigated by those low-frequency mutations that may reflect a lower fidelity of the SuperScript IV compared to the COVIDSeq reverse transcriptase. Further investigations will be needed to improve this specific step of the method. Altogether, our results confirm that MBC is compatible with NGS sequencing but can still be optimized to limit the impact of the PCR inhibitors on the quality of the genome sequences.

## 4. Discussion

Wastewater-based epidemiology (WBE) has emerged as an efficient strategy to monitor in real-time the health status of a given population, as pathogenic viruses may be present in the excrement and fluids of infected individuals. The application of WBE was previously used to monitor pathogenic viruses around the world (Lago et al., 2003, Katayama et al., 2008 and McCall et al., 2020), and recently implemented to monitor SARS-CoV-2/COVID-19. Most of the methods described in the literature have three fundamental stages for analyzing and quantifying SARS-CoV-2 virus: viral concentration, RNA extraction and quantification of target genes.

The concentration of viruses from wastewater samples is a crucial step driving the efficiency and the sensitivity of the techniques. Viral concentration methods were initially developed for analyzing and detecting non-enveloped viruses. Less is known about the recovery efficiency for enveloped viruses. There are significant structural differences between enveloped and non-enveloped viruses which could lead to differences in concentration recovery efficiencies (Wurtzer et al., 2021). Haramoto et al., 2009 demonstrated differences in virus recovery efficiencies between enveloped and non-enveloped viruses in lake water in Japan. To date, there is no standard methodology developed for the quantification and detection of SARS-CoV-2. Protocols differ according to sample volume, storage conditions, concentration method, RNA extraction method, and RNA quantification method.

To estimate SARS-CoV-2 recovery efficiencies, researchers have been using surrogate viruses such as process controls. For instance, bovine coronavirus (BCoV) was used by LaTurner et al., 2021 and Gonzalez et al., 2020, murine hepatitis virus (MHV) was used by Ahmed, Bertsch, et al., 2020, human coronavirus (HCoV-229E) was used by La Rosa et al., 2021, Phi6 was used by Sherchan et al., 2020 and recently inactivated SARS-CoV-2 was used by Zheng et al., 2023. To our knowledge, no comparison between surrogate virus recovery efficiencies and those of actual SARS-CoV-2 and between themselves were done. Thus, there is a doubt about the ability to translate recovery efficiencies of spike-in process controls to SARS-CoV-2 (Ahmed et al., 2020b). In a recent publication, Zheng and colleagues suggested that endogenous SARS-CoV-2 in wastewater samples exhibits distinct affinities to flocculants and partitioning than exogenously spiked virus. Therefore, it is advised to use positive raw wastewater samples to evaluate quantification methods and determine an optimal protocol for various applications (Zheng et al., 2023). In this study, we quantified SARS-CoV-2 and FRNAPH GGII in crude wastewater samples (C samples) and membrane-based concentrated samples (MBC). The mean recovery efficiency was 51 ± 24% and 57 ± 28% for SARS-CoV-2 and FRNAPH GGII, respectively, across 4 WWTPs demonstrating the efficiency of the MBC method to reduce the quantification limit of enveloped and non-enveloped viruses up to 100 folds for 500 mL of input. With 1 liter of input, MBC allowed to quantify SARS-CoV-2 RNA in wastewaters containing viral concentrations as low as 40 GC/L. However, large variation in viral RNA concentration efficiencies were observed within WWTPs. Considering the high repeatability of the method, these variations were not due to the technic but to the heterogeneity in the composition of the wastewater at the time of collection. Additional studies will be required to link this variability in MBC efficiency to specific water parameters. Physicochemical characteristics of the wastewater being sampled such as salinity, pH, temperature, or suspended solids will be investigated (Mohapatra et al., 2021).

The volume of wastewater input in viral concentration process may impact reproducibility, sensitivity, variability and the instrument requirements (LaTurner et al., 2021). First, increasing the volume of wastewater input may influence processing time. Indeed, a nearly exponential relationship between input volume and processing time was reported in filtration methods (LaTurner et al., 2021). With our method, we have been able to concentrate viruses from 500 mL to 1 L of wastewater sample which was much larger volumes than ones reported to date (15 mL and 50 mL), (LaTurner et al., 2021, Ahmed, Bertsch, et al., 2020) within approximate times ranging from 1 hour to 4 hours. Of note the use of multiple MBC systems allowed for the concentrations of multiple large volumes in parallel within the same time. Expanding the processed water volume will be a key area of future development for us considering the existing limitations of the technique which are determined by the membrane filtration surface and the clogging that may occur with larger volumes of highly turbidity wastewater samples.

Our method for concentrating and quantifying SARS-CoV-2 RNA is reliable and low-cost. Indeed, the consumable cost of our ultrafiltration process of wastewater is about 3 times cheaper per mL of processed water than commercial ultrafiltration methods commonly used to quantify SARS-CoV-2. In addition, our in-house RNA extraction technique (routine C method) is about 30% cheaper than RNA extractions using commercial kits. Furthermore, our routine C method does not require sophisticated and expensive equipment such as automats or high-speed centrifuges. Finally, limiting the use of commercial kits could be a crucial advantage during crisis period with limited access to kits. The routine C method presented here is brand-limited and easy to implement. It uses very common reagents and tools that can be found in most laboratories.

Previous studies highlighted the need to concentrate both solid and liquid fractions of wastewater samples (Ahmed et al., 2020b). Indeed, pre-treatment steps such as pre-filtration to remove cells and larger debris led to losses of particles associated virus in the pellet. For instance, resultant pellets from using centrifugal filter devices (Centricon Plus-70 and Amicon Ultra 15) led to 30% loss of process control (MHV) during pre-filtration step (Ahmed et al., 2020b). With our method, both solid and liquid fractions of wastewater samples were concentrated and analyzed.

Master Mix Fast Virus 1-Step TaqMan™ used in this study to quantify SARS-CoV-2 target genes is optimized to handle RT-PCR inhibitors from samples such as blood and faeces. Thereby, the co-concentration of PCR inhibitors from large volumes such as 500 mL of wastewater had no significant influence on SARS-CoV-2 quantification with our method. However, the heterogeneity of the wastewater composition and the presence of PCR inhibitors impacted the quality of the sequences obtained by NGS. We indeed observed variability in the quality of the sequences generated with different MBC samples of wastewater. Our results suggest that inhibition of the NGS enzymes is the key point to be addressed to overcome this technical issue. Moreover, we describe sequences obtained with MBC samples that harbored very good breadth of coverage and allow to identify SARS-CoV-2 mutations via the pangolin pipeline. The substitution of the COVIDSeq reverse transcriptase by the SuperScript IV further improve the coverage of the SARS-CoV-2 sequence to 100% but negatively impacted the sequence accuracy. These results suggest that COVIDSeq reverse transcriptase is more sensitive to inhibitors but have a higher fidelity. Altogether, MBC is compatible with NGS sequencing but more investigations are required to enhance the consistency of the sequencing quality, owing to the heterogeneous composition of wastewater. Wastewater is a very complex matrix, which can affect the integrity of viral particles and viral genomes to be sequenced. Future work should identify the integrity of SARS-CoV-2 genome in wastewater and how it can be affected by concentration methods. Concentration by ultrafiltration may be scaled-up (2-100L) and used for molecular detection assays. In addition, since the MBC protocol is not destructive, it can be applicable to quantification of enveloped or naked viruses, bacteria, and parasites but also to evaluate the viability of the pathogens in wastewater.

## 5. Conclusions

A new method was developed to concentrate, quantify, and sequence SARS-CoV-2 in large volumes of wastewater.

- MBC average of SARS-CoV-2 concentration efficiencies was more than 50 folds compared to the crude wastewater and up to 97 folds depending on the samples for 500 ml of input.
- MBC efficiency allows quantification of SARS-CoV-2 for concentrations below 100 and up to 40 GC/L.
- MBC does not significantly increase PCR inhibition compared to crude wastewater and is highly reproductible.
- MBC process is efficient for enveloped-viruses such as SARS-CoV-2 and naked viruses such as FRNAPH GGII.
- MBC process is suitable for wastewater-based survey of viral emergences.
- MBC is compatible with next generation sequencing but can still be improved to limit the variability of the sequence qualities due to the heterogeneity of the wastewater composition.

The routine C method presented here is easy to implement, cheaper than the commercial kits, brand limited, open-access and suitable for viral survey in wastewater.

## Declaration of competing results

The authors declare that no known competing financial interests or personal relationships could appear to influence the work reported in this paper.

## Data Availability

All data produced in the present work are contained in the manuscript

## Acknowledgments

This study was supported by the University of Strasbourg, the French scientific group of interest GIS-OBEPINE, The Agence de l’Eau Rhin Meuse, by the Grand Est region, the French national agencies for research ANR (COVIDEU project) and ANRS-MIE (EmerEaUde project). The authors would like to thank operators and local authorities from Syndicat des Eaux et de l’Assainissement Alsace-Moselle (SDEA) for the sampling efforts made in Alsace WWTPs. Special thanks to Florence Burvingt and Nathalie Caille for their support to our WWBE projects.

Work in C.V.L. lab was supported by the Belgian National Fund for Scientific Research (F.R.S.-FNRS, Belgium), the ULB-COVID-Fund and the ‘‘Les Amis des Instituts Pasteur à Bruxelles, asbl’’. The laboratory of C.V.L. is part of the ULB-Cancer Research Center (U-CRC) (Faculty of Medicine, ULB). C.V.L. is “Directrice de Recherches” of the Belgian National Fund for Scientific Research (F.R.S.-FNRS, Belgium). O.H. was a fellow of the Belgian ‘‘Fonds pour la formation à la Recherche dans l’Industrie et dans l’Agriculture (FRIA) du F.R.S.-FNRS’’ (Contract F 3/5/5 -FRIA/FC -6035) and is a fellow of ‘‘Les Amis des Instituts Pasteur à Bruxelles, asbl’’.

## Authors Contribution

G.E.S., performed experiments, analyzed data, generated figures, and contributed to the writing of the article. O.R., M.V.G., and C.W. conceived the experiments, supervised the work, analyzed the data, generated, and/or improved figures, provide fundings and wrote the article.

L.D.J. and J.V.A. performed experiments and analyzed data.

Z.G. performed NGS sequencing and analysis, contributed to scientific discussions and improved the writing. D.D., R.O., and C.D., contributed to NGS sequencing and analysis.

N.C. processed data and generated figures.

V.M. and J-F.D., contributed to the scientific discussions, improved the writing, and contributed to provide fundings.

M. B., T.L., C.S., D.T., O.H. and C.V.L. contributed to scientific discussions and improved the writing.

S.R. and D.R. managed the scientific project and the pre-analytical stage including the sampling and logistic in French Guiana.

OBEPINE consortium supports French WWBE projects, organizes the logistic of WW sampling, contributes to global scientific orientations and contributes to provide fundings.

## OBEPINE consortium associated members

Maday Y., Sorbonne Université, CNRS, Université Paris Cité, Laboratoire Jacques-Louis Lions (LJLL), UMR 7598 CNRS, Paris, France. Mouchel JM. from Sorbonne University, CNRS, EPHE, UMR 7619 Metis, Paris, France; Boudaud N., from Actalia, Food Safety Department, F-50000 Saint-Lô, France; Gantzer C. and Bertrand I. from LCPME, UMR 7564, CNRS, Université de Lorraine, F-54000 Nancy, France; Peyrefitte C^6^, Raffestin S^6^, Rousset D^6^; Boni M^8^; Marechal V^5^ ; Wallet C^1^ and Rohr O^1^.

## References

Ahmed, W., Angel, N., Edson, J., Bibby, K., Bivins, A., O’Brien, J.W., Choi, P.M., Kitajima, M., Simpson, S.L., Li, J., Tscharke, B., Verhagen, R., Smith, W.J.M., Zaugg, J., Dierens, L., Hugenholtz, P., Thomas, K.V., Mueller, J.F., 2020a. First confirmed detection of SARS-CoV-2 in untreated wastewater in Australia: A proof of concept for the wastewater surveillance of COVID-19 in the community. Sci Total Environ 728, 138764. 10.1016/j.scitotenv.2020.138764

Ahmed, W., Bertsch, P.M., Bivins, A., Bibby, K., Farkas, K., Gathercole, A., Haramoto, E., Gyawali, P., Korajkic, A., McMinn, B.R., Mueller, J.F., Simpson, S.L., Smith, W.J.M., Symonds, E.M., Thomas, K.V., Verhagen, R., Kitajima, M., 2020b. Comparison of virus concentration methods for the RT-qPCR-based recovery of murine hepatitis virus, a surrogate for SARS-CoV-2 from untreated wastewater. Science of The Total Environment 739, 139960. 10.1016/j.scitotenv.2020.139960

Bertrand, I., Challant, J., Jeulin, H., Hartard, C., Mathieu, L., Lopez, S., Scientific Interest Group Obépine Schvoerer, E., Courtois, S., Gantzer, C., 2021. Epidemiological surveillance of SARS-CoV-2 by genome quantification in wastewater applied to a city in the northeast of France: Comparison of ultrafiltration- and protein precipitation-based methods. Int J Hyg Environ Health 233, 113692. 10.1016/j.ijheh.2021.113692

Bioanalytical Method Validation. Guidance for Industry, 2018. . prepared by the Biopharmaceutics Coordinating Committee of the US Dept. of Health and Human Services Food and Drug Administration/ Center for Drug Evaluation (CDER) in cooperation with the Center for Veterinary Medicine (CVM).

Corman, V.M., Landt, O., Kaiser, M., Molenkamp, R., Meijer, A., Chu, D.K., Bleicker, T., Brünink, S., Schneider, J., Schmidt, M.L., Mulders, D.G., Haagmans, B.L., van der Veer, B., van den Brink, S., Wijsman, L., Goderski, G., Romette, J.-L., Ellis, J., Zambon, M., Peiris, M., Goossens, H., Reusken, C., Koopmans, M.P., Drosten, C., 2020. Detection of 2019 novel coronavirus (2019-nCoV) by real-time RT-PCR. Euro Surveill 25, 2000045. 10.2807/1560-7917.ES.2020.25.3.2000045

Euliano, E.M., Hardcastle, A.N., Victoriano, C.M., Gabella, W.E., Haselton, F.R., Adams, N.M., 2019. Multiplexed Adaptive RT-PCR Based on L-DNA Hybridization Monitoring for the Detection of Zika, Dengue, and Chikungunya RNA. Sci Rep 9, 11372. 10.1038/s41598-019-47862-6

Gerber, Z., Daviaud, C., Delafoy, D., Sandron, F., Alidjinou, E.K., Mercier, J., Gerber, S., Meyer, V., Boland, A., Bocket, L., Olaso, R., Deleuze, J.-F., 2022. A comparison of high-throughput SARS-CoV-2 sequencing methods from nasopharyngeal samples. Sci Rep 12, 12561. 10.1038/s41598-022-16549-w

Gonzalez, R., Curtis, K., Bivins, A., Bibby, K., Weir, M.H., Yetka, K., Thompson, H., Keeling, D., Mitchell, J., Gonzalez, D., 2020. COVID-19 surveillance in Southeastern Virginia using wastewater-based epidemiology. Water Research 186, 116296. 10.1016/j.watres.2020.116296

Haramoto, E., Kitajima, M., Katayama, H., Ito, T., Ohgaki, S., 2009. Development of virus concentration methods for detection of koi herpesvirus in water. Journal of fish diseases 32, 297–300. 10.1111/j.1365-2761.2008.00977.x

Katayama, H., Haramoto, E., Oguma, K., Yamashita, H., Tajima, A., Nakajima, H., Ohgaki, S., 2008. One-year monthly quantitative survey of noroviruses, enteroviruses, and adenoviruses in wastewater collected from six plants in Japan. Water Research 42, 1441–1448. 10.1016/j.watres.2007.10.029

Kitajima, M., Ahmed, W., Bibby, K., Carducci, A., Gerba, C.P., Hamilton, K.A., Haramoto, E., Rose, J.B., 2020. SARS-CoV-2 in wastewater: State of the knowledge and research needs. Science of The Total Environment 739, 139076. 10.1016/j.scitotenv.2020.139076

La Rosa, G., Mancini, P., Bonanno Ferraro, G., Veneri, C., Iaconelli, M., Bonadonna, L., Lucentini, L., Suffredini, E., 2021. SARS-CoV-2 has been circulating in northern Italy since December 2019: Evidence from environmental monitoring. Science of The Total Environment 750, 141711. 10.1016/j.scitotenv.2020.141711

Lago, P.M., Gary, H.E., Jr, Pérez, L.S., Cáceres, V., Olivera, J.B., Puentes, R.P., Corredor, M.B., Jímenez, P., Pallansch, M.A., Cruz, R.G., 2003. Poliovirus detection in wastewater and stools following an immunization campaign in Havana, Cuba. International Journal of Epidemiology 32, 772–777. 10.1093/ije/dyg185

LaTurner, Z.W., Zong, D.M., Kalvapalle, P., Gamas, K.R., Terwilliger, A., Crosby, T., Ali, P., Avadhanula, V., Santos, H.H., Weesner, K., Hopkins, L., Piedra, P.A., Maresso, A.W., Stadler, L.B., 2021. Evaluating recovery, cost, and throughput of different concentration methods for SARS-CoV-2 wastewater-based epidemiology. Water Research 197, 117043. 10.1016/j.watres.2021.117043

Maréchal, V., Maday, Y., Wallet, C., Cluzel, N., Borde, C., 2023. Wastewater-based epidemiology: Retrospective, current status, and future prospects. Anaesthesia Critical Care & Pain Medicine 42, 101251. 10.1016/j.accpm.2023.101251

McCall, C., Wu, H., Miyani, B., Xagoraraki, I., 2020. Identification of multiple potential viral diseases in a large urban center using wastewater surveillance. Water Research 184, 116160. 10.1016/j.watres.2020.116160

McMinn, B.R., Korajkic, A., Kelleher, J., Herrmann, M.P., Pemberton, A.C., Ahmed, W., Villegas, E.N., Oshima, K., 2021. Development of a large volume concentration method for recovery of coronavirus from wastewater. Sci Total Environ 774, 145727. 10.1016/j.scitotenv.2021.145727

Medema, G., Been, F., Heijnen, L., Petterson, S., 2020a. Implementation of environmental surveillance for SARS-CoV-2 virus to support public health decisions: Opportunities and challenges. Curr Opin Environ Sci Health 17, 49–71. 10.1016/j.coesh.2020.09.006

Medema, G., Heijnen, L., Elsinga, G., Italiaander, R., Brouwer, A., 2020b. Presence of SARS-Coronavirus-2 in sewage. 10.1101/2020.03.29.20045880

Mohapatra, S., Menon, N. Gayathri Mohapatra, G., Pisharody, L., Pattnaik, A., Menon, N. Gowri, Bhukya, P.L., Srivastava, M., Singh, M., Barman, M.K., Gin, K.Y.-H., Mukherji, S., 2021. The novel SARS-CoV-2 pandemic: Possible environmental transmission, detection, persistence and fate during wastewater and water treatment. Science of The Total Environment 765, 142746. 10.1016/j.scitotenv.2020.142746

O’Toole, Á., Scher, E., Underwood, A., Jackson, B., Hill, V., McCrone, J.T., Colquhoun, R., Ruis, C., Abu-Dahab, K.,Taylor, B., Yeats, C., du Plessis, L., Maloney, D., Medd, N., Attwood, S.W., Aanensen, D.M., Holmes, E.C., Pybus, O.G., Rambaut, A., 2021. Assignment of epidemiological lineages in an emerging pandemic using the pangolin tool. Virus Evol 7, veab064. 10.1093/ve/veab064

Peccia, J., Zulli, A., Brackney, D.E., Grubaugh, N.D., Kaplan, E.H., Casanovas-Massana, A., Ko, A.I., Malik, A.A., Wang, D., Wang, M., Warren, J.L., Weinberger, D.M., Omer, S.B., 2020. SARS-CoV-2 RNA concentrations in primary municipal sewage sludge as a leading indicator of COVID-19 outbreak dynamics. 10.1101/2020.05.19.20105999

Sherchan, S.P., Shahin, S., Ward, L.M., Tandukar, S., Aw, T.G., Schmitz, B., Ahmed, W., Kitajima, M., 2020. First detection of SARS-CoV-2 RNA in wastewater in North America: A study in Louisiana, USA. Sci Total Environ 743, 140621. 10.1016/j.scitotenv.2020.140621

Wolf, S., Hewitt, J., Greening, G.E., 2010. Viral Multiplex Quantitative PCR Assays for Tracking Sources of Fecal Contamination. Appl Environ Microbiol 76, 1388–1394. 10.1128/AEM.02249-09

Wurtzer, S., Marechal, V., Mouchel, J.M., Maday, Y., Teyssou, R., Richard, E., Almayrac, J.L., Moulin, L., 2020. Evaluation of lockdown effect on SARS-CoV-2 dynamics through viral genome quantification in waste water, Greater Paris, France, 5 March to 23 April 2020. Euro Surveill 25, 2000776. 10.2807/1560-7917.ES.2020.25.50.2000776

Wurtzer, S., Waldman, P., Ferrier-Rembert, A., Frenois-Veyrat, G., Mouchel, J.M., Boni, M., Maday, Y., OBEPINE consortium, Marechal, V., Moulin, L., 2021. Several forms of SARS-CoV-2 RNA can be detected in wastewaters: Implication for wastewater-based epidemiology and risk assessment. Water Res 198, 117183. 10.1016/j.watres.2021.117183

Xiao, F., Sun, J., Xu, Y., Li, F., Huang, X., Li, H., Zhao, Jingxian, Huang, J., Zhao, Jincun, 2020. Infectious SARS-CoV-2 in Feces of Patient with Severe COVID-19. Emerg Infect Dis 26, 1920–1922. 10.3201/eid2608.200681

Zheng, X., Wang, M., Deng, Y., Xu, X., Lin, D., Zhang, Y., Li, S., Ding, J., Shi, X., Yau, C.I., Poon, L.L.M., Zhang, T., 2023. A rapid, high-throughput, and sensitive PEG-precipitation method for SARS-CoV-2 wastewater surveillance. Water Research 230, 119560. 10.1016/j.watres.2022.119560

