## supplemental figures for "Highly efficient and sensitive membrane-based concentration process allows quantification, surveillance, and sequencing of viruses in large volumes of wastewater"

***Supplementary Table 1. Characteristics of the different wastewater treatment systems.***

| Wastewater Treatment Plant (Anonymized) | Nominal capacity (population equivalent) | Wastewater flow rate ($m^3$/day) | COD (kg/day) | pH | Conductivity (µS/cm) | OD (600 nm) |
|---|---|---|---|---|---|---|
| WWTP1 | 30 000 | 3573 | 1616 | 7.35 | 1317 | 0.649 |
| WWTP2 | 35 000 | 3273 | 570 | 7.44 | 668 | 0.289 |
| WWTP3 | 25 000 | 1347 | 311 | 6.92 | 579 | 0.370 |
| WWTP4 | 21 000 | 3308 | 824 | 7.40 | 1069 | 0.405 |

Population equivalent, wastewater flow rate and chemical oxygen demand (COD) were provided by the Syndicat des Eaux et de l'Assainissement d'Alsace-Moselle (SDEA). pH, Conductivity and Optical Density (OD) were measured (average value).

***Supplementary Table 2. Primers used for RT-qPCR quantifications.***

| Oligo Name | MW [g/mol] | GC - Content | Tm | Sequence (5'->3') | Length [mer] | 5' Mod. | 3' Mod. |
|---|---|---|---|---|---|---|---|
| E_Sarbeco_F | 8033 | 34 | 58.5 | ACAGGTACGTTAATAGTTAATAGCGT | 26 | | |
| E_Sarbeco_R | 6712 | 45 | 58.4 | ATATTGCAGCAGTACGCACACA | 22 | | |
| E_Sarbeco_P1 | 8934 | 53 | 66.4 | [FAM]ACACTAGCCATCCTTACTGCGCTTCG[BHQ1] | 26 | FAM | BHQ1 |
| Dengue Fw | 6176 | 42 | 54.2 | CAAAAGGAAGTCGYGCAATA | 20 | | |
| Dengue Rv | 7311 | 43 | 60.2 | CTGAGTGAATTCTCTCTGCTRAAC | 24 | | |
| Dengue-Probe | 6977 | 68 | 63.1 | [FAM]CATGTGGYTGGGAGCRCGC[BHQ1] | 19 | FAM | BHQ1 |
| VTB4-FphGIIf | 6714 | 45 | 58.4 | ACCTATGTTCCGATTCASAGAG | 22 | | |
| VTB4-FphGIIr | 6479 | 47 | 57.9 | GGTAGGCAAGTCCATCAAAGT | 21 | | |
| VTB4-FphGIIprobe | 8403 | 54 | 64.4 | [FAM]CACTCGCGATTGTGCTGTCCGATT[BHQ1] | 24 | FAM | BHQ1 |
| nCoV_IP4-14059Fw | 5874 | 42 | 52.4 | GGTAACTGGTATGATTTCG | 19 | | |
| nCoV_IP4-14146Rv | 6196 | 40 | 53.2 | CTGGTCAAGGTTAATATAGG | 20 | | |
| nCoV_IP4-14084Probe(+) | 6843 | 52 | 56.7 | [FAM]TCATACAAACCACGCCAGG[BHQ1] | 19 | FAM | BHQ1 |

*Supplementary Figure 1*

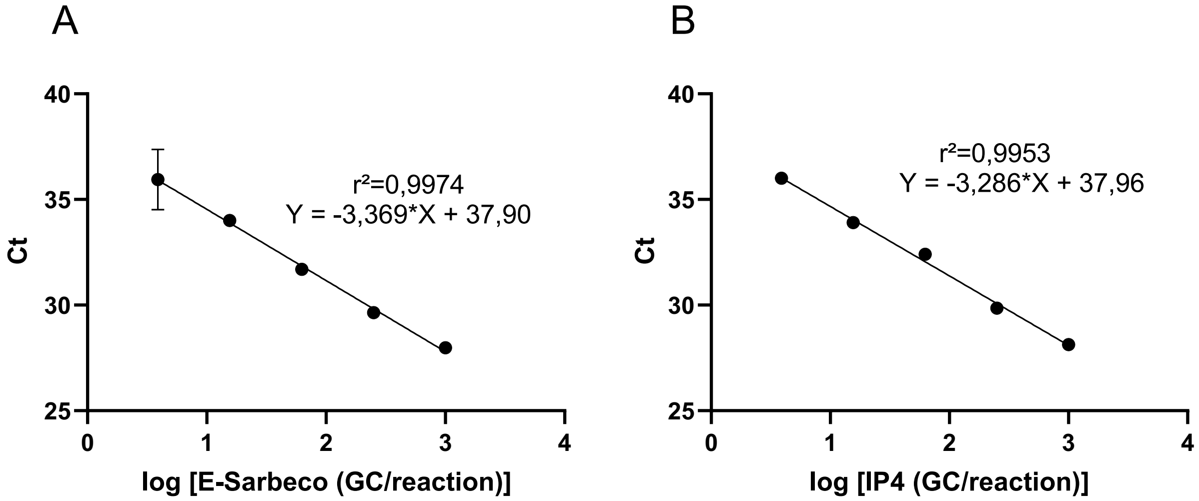


***Standard curve of SARS-CoV-2 E-Sarbeco (E gene) and IP4 (RdRp gene) (A. and B. respectively).***
Quantified SARS-CoV-2 RNA was diluted in a fourfold dilution series from 1000 genome copies/reaction to 1 genome copy/reaction. Cp for each dilution point was determined by qPCR in 2 replicates respectively. Standard curves have been routinely repeated. A representative standard curve for each target is presented.

E-Sarbeco and RdRp-IP4 standard curves had a dynamic linear range of quantification from 1 to $10^3$ genome copies/reaction (Fig. 3). The LoD were determined at 15 genome copies/reaction for a Ct value of 34.59 ± 0.46. Detection was possible until 3.9 genome copies/reaction, and linearity was conserved until this point. The LoD for the IP4 quantification was determined at 15 genomes copies/reaction for a Ct value of 34.77 ± 0.81. Detection was possible until 3.9 genome copies/reaction, and linearity was conserved until this point.

*Supplementary Figure 2*

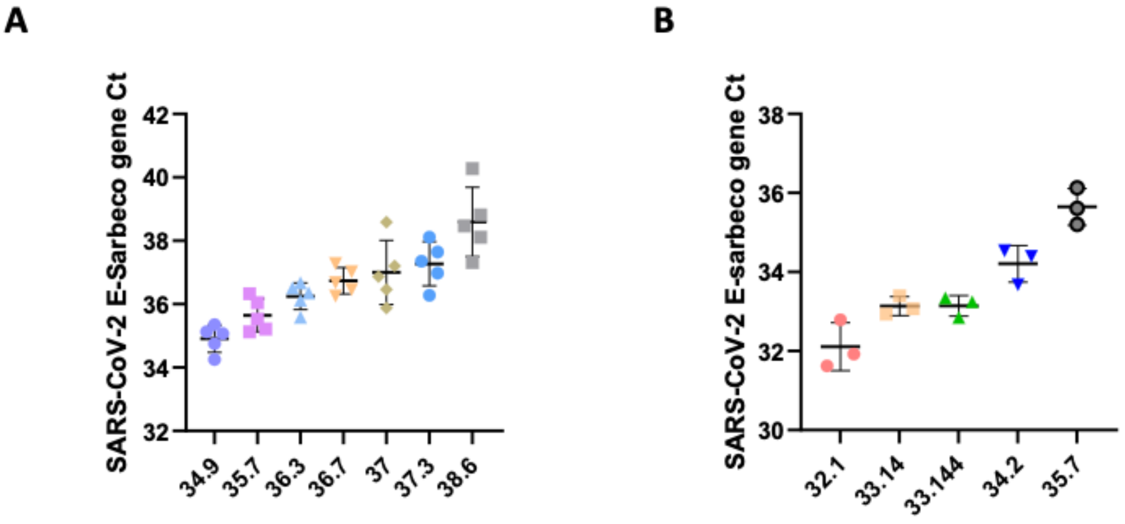


**Repeatability of SARS-CoV-2 quantification method using crude wastewater samples.**
A: Ct of SARS-CoV-2 target E-Sarbeco gene for 7 different crude wastewater samples extracted in replicates (n=5), plotted against the corresponding mean. B: Ct of SARS-CoV-2 target E-Sarbeco gene for 5 different inter-laboratory samples extracted in triplicate, plotted against the corresponding mean. Means and standard deviations are shown on these figures.

To determine the intra-assay precision of our SARS-CoV-2 quantification method from crude wastewater samples (C samples), 7 different samples were analyzed in 5 replicates. Results from RNA extraction and quantification replicates were plotted against the corresponding mean (Supp Fig A). The relative standard deviation (R.S.D.) to the mean ranged from 1.2 to 2.8%.
To complete this first set of experiments, 5 other samples from inter-laboratory essays were analyzed in triplicates. SARS-CoV-2 E-Sarbeco gene Ct for inter-laboratory samples quantified by our laboratory were plotted against the corresponding mean (Supp Fig. 1B). The R.S.D. for samples with SARS-CoV-2 mean Ct ranged from 0.7 to 1.9%.
The repeatability can be expressed as the R.S.D. (to the mean), which was on average 1.5% for all samples analyzed. This R.S.D of 1.5% is far below the acceptable limit of 15% as described by Biopharmaceutics Coordinating Committee of the US Food and Drug Administration ("Bioanalytical Method Validation. Guidance for Industry," 2018). Thus, our results demonstrated the high repeatability of our SARS-CoV-2 extraction and quantification methods.

*Supplementary Figure 3*

**A**

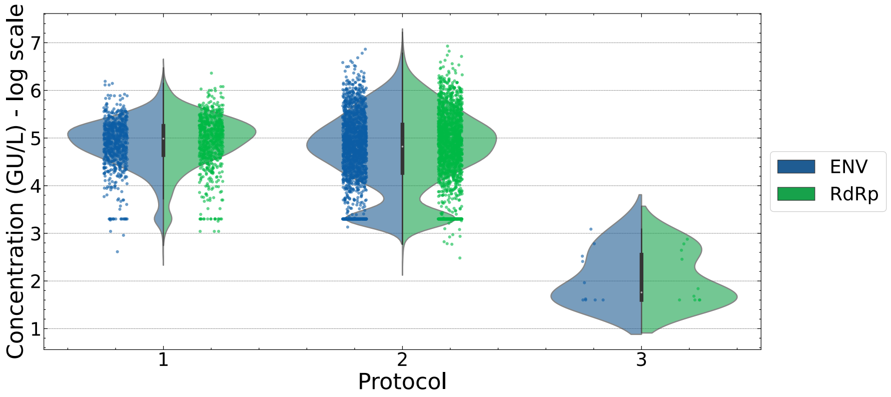


**B**

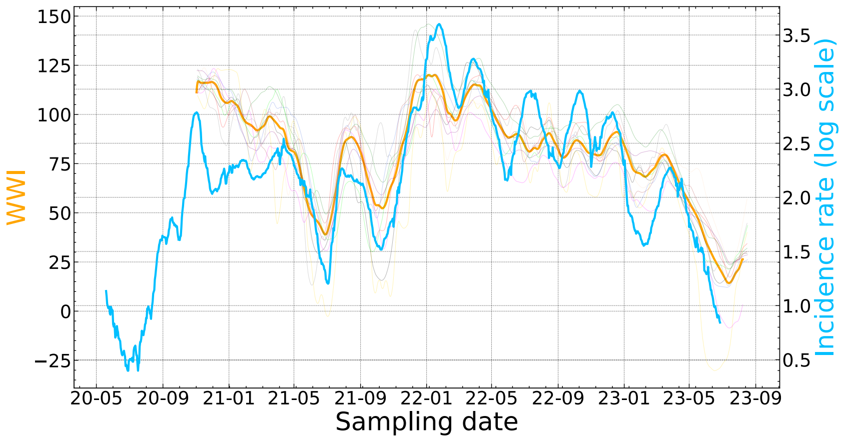


**The routine C and MBC methods are suitable for SARS-CoV2 surveillance in wastewater.**

(A) Quantifications of SARS-CoV-2 ENV (Blue) and SARS-CoV-2 RdRp (Green) genes during the weekly based SARS-CoV-2 survey in different WWTP with the NucliSens® EasyMAG$^{TM}$ method (1), the routine C method (2) and the MBC method (3) are presented. (B) The OBEPINE WasteWater Indicator (WWI) corresponding to the 11 WWTPs in Alsace- France (yellow) is presented with the incidence rate of the same region.

As shown in panel A, the routine C method (protocol 2) is comparable to the NucliSens® EasyMAG$^{TM}$ kit (protocol 1). Of note a significant number of quantifications gave results close to the theoretical LOD (bars) confirming that the effective LOD is close to the theoretical once. Some samples that had undetectable genome copies with the routine C method were subjected to the MBC process (protocol 3). As shown, we were able to detect SARS-CoV-2 concentrations very close to the theoretical LOD that is set between 20 and 40 GC/L depending on the volume of wastewater processed (1 L or 500mL).

A shown in panel B, a strong correlation between the incidence rate and the OBEPINE indicator is observed (Pearson = 0.8) confirming the accuracy of the routine C protocol to monitor SARS-CoV-2 circulation in a large population.
